# Cell-density independent increased lymphocyte production and loss rates post-autologous hematopoietic stem cell transplantation

**DOI:** 10.1101/2020.04.24.20078295

**Authors:** Mariona Baliu-Piqué, Vera van Hoeven, Julia Drylewicz, Lotte E. van der Wagen, Anke Janssen, Sigrid A. Otto, Menno C. van Zelm, Rob J. de Boer, Jürgen Kuball, José A.M. Borghans, Kiki Tesselaar

**Affiliations:** Center for Translational Immunology, University Medical Center Utrecht, The Netherlands; Department of Experimental Immunology, Amsterdam UMC, University of Amsterdam, The Netherlands; Department of Hematology, University Medical Center Utrecht, The Netherlands; Department of Immunology and Pathology, Monash University and Alfred Hospital, Melbourne; Theoretical Biology, Utrecht University, Utrecht, The Netherlands

## Abstract

Lymphocyte numbers need to be quite tightly regulated. It is generally assumed that lymphocyte production and survival rates increase homeostatically when lymphocyte numbers decrease. This widely-accepted concept is largely based on experiments in mice. In humans, lymphocyte reconstitution usually occurs very slowly, which challenges the idea that density dependent homeostasis aids recovery from lymphopenia. Using *in vivo* deuterium labelling, we quantified lymphocyte production and survival rates in patients who underwent an autologous hematopoietic stem cell transplantation (autoHSCT). We indeed found that the production rates of most T-cell and B-cell subsets in autoHSCT-patients were 2 to 8-times higher than in healthy controls. These increased lymphocyte production rates went hand in hand with a 3 to 9-fold increase in cell loss rates, and both rates did not normalize when cell numbers did. This challenges the concept of homeostatic regulation of lymphocyte production and survival rates in humans.

## Introduction

Under healthy conditions, the peripheral T- and B-cell populations are maintained at relatively stable numbers throughout life ^1,2^. Homeostatic mechanisms are thought to regulate lymphocyte production and survival rates in a density-dependent manner. Indeed, studies in rodents have shown that lymphocyte division and survival rates increase in response to severe lymphopenic conditions ^3^. Robust peripheral proliferation of T-cells occurs both upon adoptive cell transfer into severely lymphocyte-depleted mice, and in partially immune-depleted hosts in the absence of adoptive cell transfer, a phenomenon termed lymphopenia-induced proliferation (LIP) ^3–6^. Similarly, rapid proliferation and extended survival of B-cells occur after adoptive cell transfer into B-cell deficient hosts and correlate with peripheral B-cell numbers ^7^.

In analogy, it is generally assumed that lymphopenic conditions induce alterations in lymphocyte dynamics in humans. However, in humans full recovery of the T-cell compartment following an autologous hematopoietic stem cell (autoHSCT) is notoriously slow, often taking several years ^8–11^. On the basis of elevated frequencies of Ki-67^+^ cells, severe lymphopenia arising after HSCT and lymphocyte-depleting treatments has been associated with increased proliferation of naive and memory T-cells ^12–15^. However, elevated frequencies of Ki-67^+^ cells were shown to decline within 3 to 6 months after cell depletion, despite the fact that patients were still deeply lymphopenic ^13–15^. Furthermore, increased T-cell proliferation rates after allogeneic HSCT have been shown to correlate with the occurrence of graft-versus-host disease (GVHD) and infectious-disease-related complications ^14^. Together, these observations question whether homeostatic mechanisms are induced to compensate for low lymphocyte numbers in humans undergoing HSCT. It remains unclear to what extent increased T-cell proliferation post-HSCT reflects a T-cell density-dependent response to lymphopenia, or an immune response triggered by therapy-related tissue damage, infectious complications, or immune activation.

To elucidate whether lymphocyte production and death rates in humans are regulated in a density-dependent manner, we used *in vivo* deuterium labelling to quantify the production and loss rates of different T- and B-cell subsets in patients who received an autologous HSCT (autoHSCT), and had no signs of clinically manifested infections or GVHD. Twelve months after autoHSCT, absolute numbers of CD4^+^ T-cells and memory and natural effector B-cells in these patients were still lower than in healthy individuals, while CD8^+^ T-cell and naive B-cell numbers had already recovered to healthy control values. Deuterium labelling revealed that the production rates of most lymphocyte subsets, even those that had already reconstituted, were significantly higher in patients post-autoHSCT than in healthy individuals. These increased rates of T- and B-cell production could only be reconciled with the observed slow changes in lymphocyte numbers over time if lymphocyte loss rates were also significantly increased, impeding a timely reconstitution of the lymphocyte pool post-autoHSCT.

## Materials and Methods

Complete Materials and Methods can be found as Supplementary Information.

### Patient characteristics

Six patients who received an autoHSCT for the treatment of a hematologic malignancy were enrolled in the study after having provided written informed consent. Following repeated subcutaneous injections with granulocyte-colony stimulating factor (G-CSF), stem cells were obtained by leukapheresis of peripheral blood. Patients received a non T-cell depleted graft; the average number of CD34^+^ cells transplanted was 5.03×10^6^ cells/kg (median, 4.12; range, 1.82-12.38). Patients were included in the study between 196 and 420 days after autoHSCT, and had no signs of transplantation-related complications, severe infections (HIV, HBV, HCV), other liver disease, active uncontrolled infections (such as infectious mononucleosis), inadequate liver or kidney function, or cardiovascular disease before and during the study. Additional inclusion criteria were: fully transfusion-independent at start of the study, hemoglobin level ≥6mmol/l, and platelet count ≥50×10^9^/L. Any use of medication during the study was unrelated to the malignancy and the HSCT (Figure 1). In order to compare the phenotypes of the B- and T-cell compartments of patients to those of age-matched healthy individuals, we used data from healthy individuals from a previous study ^16^, and additional blood samples were collected from healthy volunteers not following the labelling protocol after having provided informed consent. This study was approved by the medical ethical committee of the University Medical Center Utrecht and conducted in accordance with the Helsinki Declaration.

**Figure 1.**
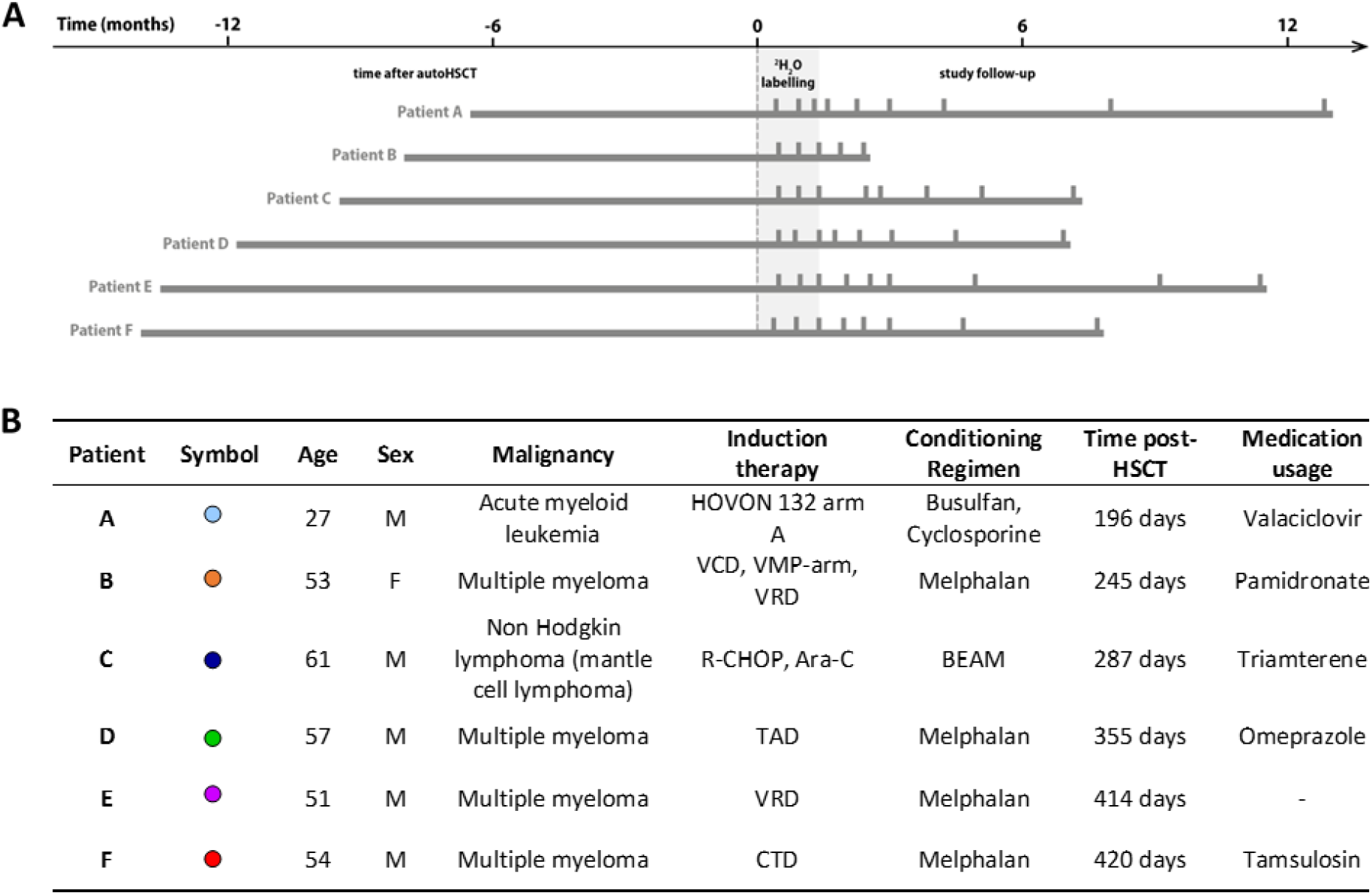
Study protocol timeline and patient characteristics. **(A)** Summary figure depicting the study time line of every patient. Patients are centred by start of ^2^H_2_O labelling. The left bar indicated the time between the autologous hematopoietic stem cell transplantation (autoHSCT) and the start of the labelling period, the grey area indicates the 6 weeks ^2^H_2_O labelling period, the right bar provides the follow up period and the vertical bars indicate the blood sampling time points. **(B)** Patient characteristics. Age=Age at start ^2^H_2_O labelling; M=Male; F=Female; Time post-HSCT=Reconstitution period at start ^2^H_2_O labelling; Medication usage=Medication during the study; HOVON 132 arm A (Idarubicin, Ara-C (Cytarabine), Daunorubicin); VCD (Bortezomib, Cyclophosphamide, Dexamethason); VMP (Bortezomib, Melphalan, Prednisone); VRD (Bortezomib, lenalidomide, dexamethasone); R-CHOP (Rituximab, Cyclophosphamide, Adriamycin, Vincristin, Prednisone); Ara-C (Cytarabine), TAD (Thalidomide, Adriamycin,Dexamethasone); CTD (Carfilzomib, Thalidomide, Dexamethasone); BEAM (Carmustine, Etoposide, Ara-C (Cytarabine), Melphalan). For absolute leukocytes, neutrophils, lymphocytes and monocytes numbers see Sup. Figure 6.

### *In vivo* deuterium labelling

*In vivo* deuterium labelling was performed as previously described with small adaptations ^16^. Briefly, patients received an oral ramp-up dose of 7.5 ml of heavy water (^2^H_2_O, 99.8% enriched, Cambridge Isotope Laboratories) per kg body water on the first day of the study, and drank a daily maintenance dose of 1.25 ml ^2^H_2_O per kg body water for 6 weeks. To reduce the study burden, the labelling period of patients was 3 weeks shorter than the previously used labelling period for healthy individuals.

### Cell isolation, flow cytometry and cell sorting

Peripheral blood mononuclear cells were obtained by Ficoll-Paque (GE Healthcare, Little Chalfont, UK) density gradient centrifugation from heparinized blood. Granulocytes were obtained by 2 cycles of erythrocyte lysis (155mM NH4Cl, 10mM KHCO3, 0.1mM Na2-EDTA, pH=7.0) of the granulocyte/erythrocyte layer. To determine baseline enrichment, total peripheral blood mononuclear cells were frozen on the first day of the study, prior to ^2^H_2_O intake.

Absolute cell numbers were determined using TruCount tubes (BD Biosciences, San Jose, CA, USA), and cell cycle analysis was performed by measuring expression of Ki-67. Samples were analysed on an LSR-II or LSR-Fortessa flow cytometer using FACS Diva software (BD Biosciences).

CD19^+^ naive (IgM^+^CD27^-^), Ig class-switched (IgM^-^CD27^+^) and IgM^+^ (IgM^+^CD27^+^) memory B-cells and CD3^+^CD4^+^ and CD3^+^CD8^+^ naive (CD27^+^CD45RO^-^) and memory (CD45RO^+^) T-cells were sorted on a FACSAria II or FACSAria III cell sorter using FACS Diva software (BD Biosciences). Flow cytometric analyses and cell sorting were always performed on freshly isolated material. Representative density dot plots and the gating strategy for TruCount analysis and cell sorting are shown in Sup. Figure 1.

### Statistical analyses

Medians were compared between groups using Mann–Whitney tests. Correlations were analysed using the Spearman’s rank test (GraphPad Software, Inc). Differences with a p-value<0.05 were considered significant.

## Results

### Heterogeneous T-cell reconstitution kinetics post-autoHSCT

To investigate whether lymphocyte production and loss depend on cell numbers during lymphopenia in humans, we quantified the production and loss rates of B- and T-cells in 6 patients who received an autoHSCT for the treatment of haematological malignancies. Patients were included in the study between 196 days and 420 days post-autoHSCT, they received deuterated water (^2^H_2_O) for six weeks, and were followed for approximately one year after start of the labelling period (Figure 1). *Patient B* withdrew from the study 10 weeks after the start of ^2^H_2_O labelling due to infectious complications unrelated to participation in the study. All other patients had no complications that needed treatment during the study follow-up, which was supported by CRP levels in the normal range (Sup. Figure 2).

**Figure 2.**
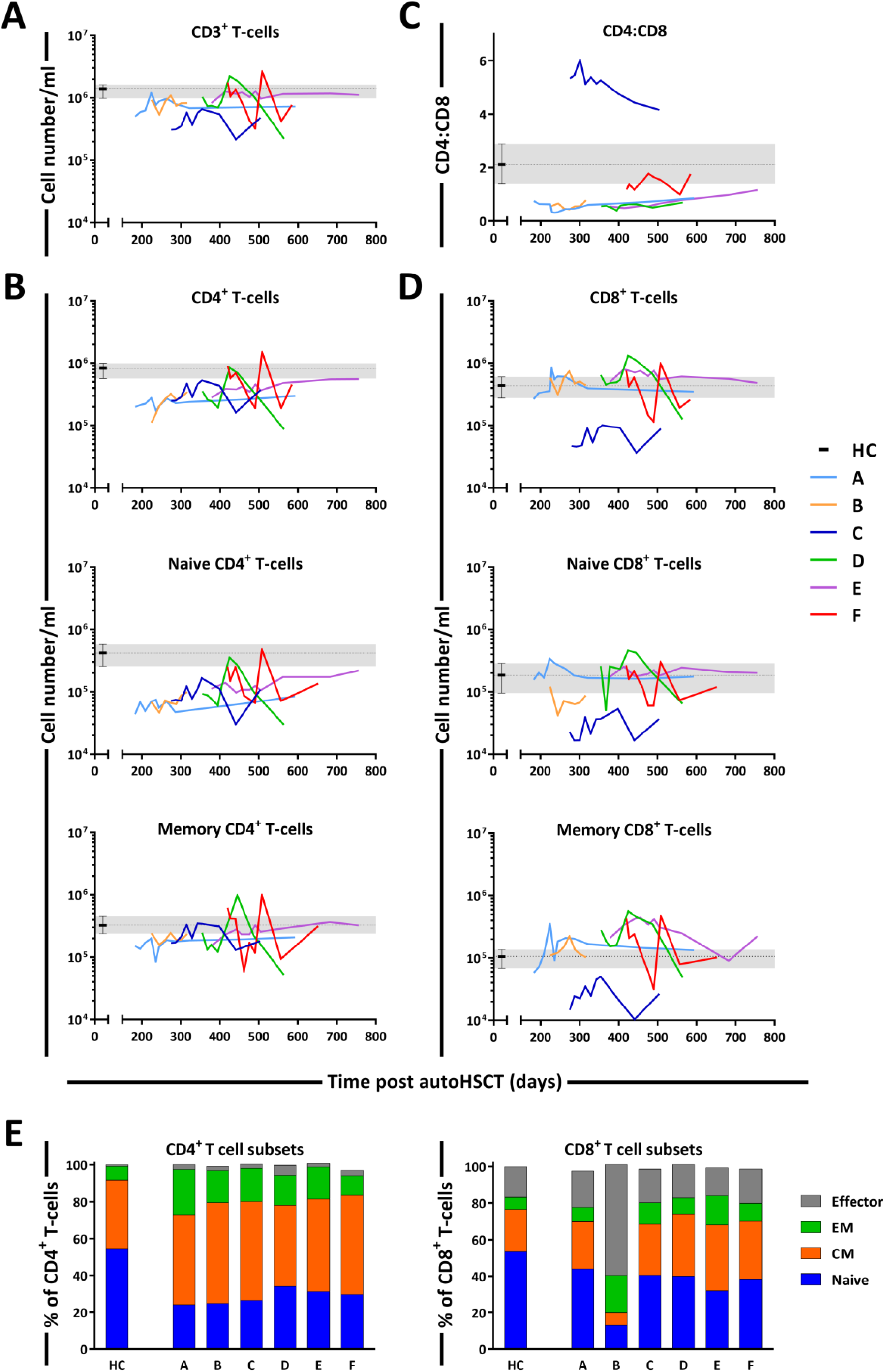
T-cell reconstitution following autoHSCT. **(A)** Absolute numbers (cells per milliliter) of total CD3^+^ T-cells. **(B)** Absolute numbers (cells per milliliter) of CD4^+^ T-cells (CD3^+^CD4^+^), naive (CD27^+^CD45RO^-^) and memory (CD45RO^+^) CD4^+^ T-cells. **(C)** CD4:CD8 ratio. **(D)** Absolute numbers (cells per milliliter) of CD8^+^ T-cells (CD3^+^CD8^+^), naive (CD27^+^CD45RO^-^) and memory (CD45RO^+^) CD8^+^ T-cells. **(A-D)** Graphs show the absolute cell counts per milliliter or the CD4:CD8 ratio in autoHSCT patients (patients A to F) over time from for the duration of the study, and the median and the corresponding IR for HCs (healthy controls, grey area and whisker bars). **(E)** Bar graphs show the median percentage of naive (CD27^+^CD45RO^-^), central memory (CM, CD27^-^CD45RO^+^), effector memory (EM, CD27^+^CD45RO+) and effector (CD27^-^CD45RO^-^) CD4^+^ and CD8^+^ T-cells of autoHSCT patients (patients A to F) and HCs (naive in blue, CM in orange, EM in green and effector in grey). For patient characteristics see Figure 1B, for the T-cell subset distribution per patient over time see Sup. Figure 3.

The sub-optimal T-cell recovery observed in the peripheral blood of patients post auto-HSCT (Figure 2A) was largely due to the slow reconstitution of CD4^+^ T-cells (Figure 2B). At the start of ^2^H_2_O labelling, CD8^+^ T-cell numbers had reached normal levels in most patients, whereas CD4^+^ T-cell numbers remained below normal levels even 1.5 years post-autoHSCT. This resulted in an inverse CD4:CD8 ratio in all patients except for *patient C* (Figure 2C), who experienced extremely slow CD8^+^ T-cell reconstitution (Figure 2D). Naive (CD45RO^-^CD27^+^) CD4^+^ T-cell numbers remained below normal levels throughout the 2-year follow-up period, whereas memory (CD45RO^+^) CD4^+^ T-cells reached the lower range of normal levels around 400 days post-autoHSCT (Figure 2B). Naive and memory CD8^+^ T-cell numbers were at normal or supra-normal levels at the start of the study in all patients except for *patient C* (Figure 2D). In line with cell numbers, for most patients the fractions of naive cells, central memory (CM, CD45RO^+^CD27^+^), effector memory (EM, CD45RO^+^CD27^-^) and effector (CD45RO^-^CD27^-^) T-cells differed from those in healthy controls and varied slightly over time (Figure 2E and Sup. Figure 3).

**Figure 3.**
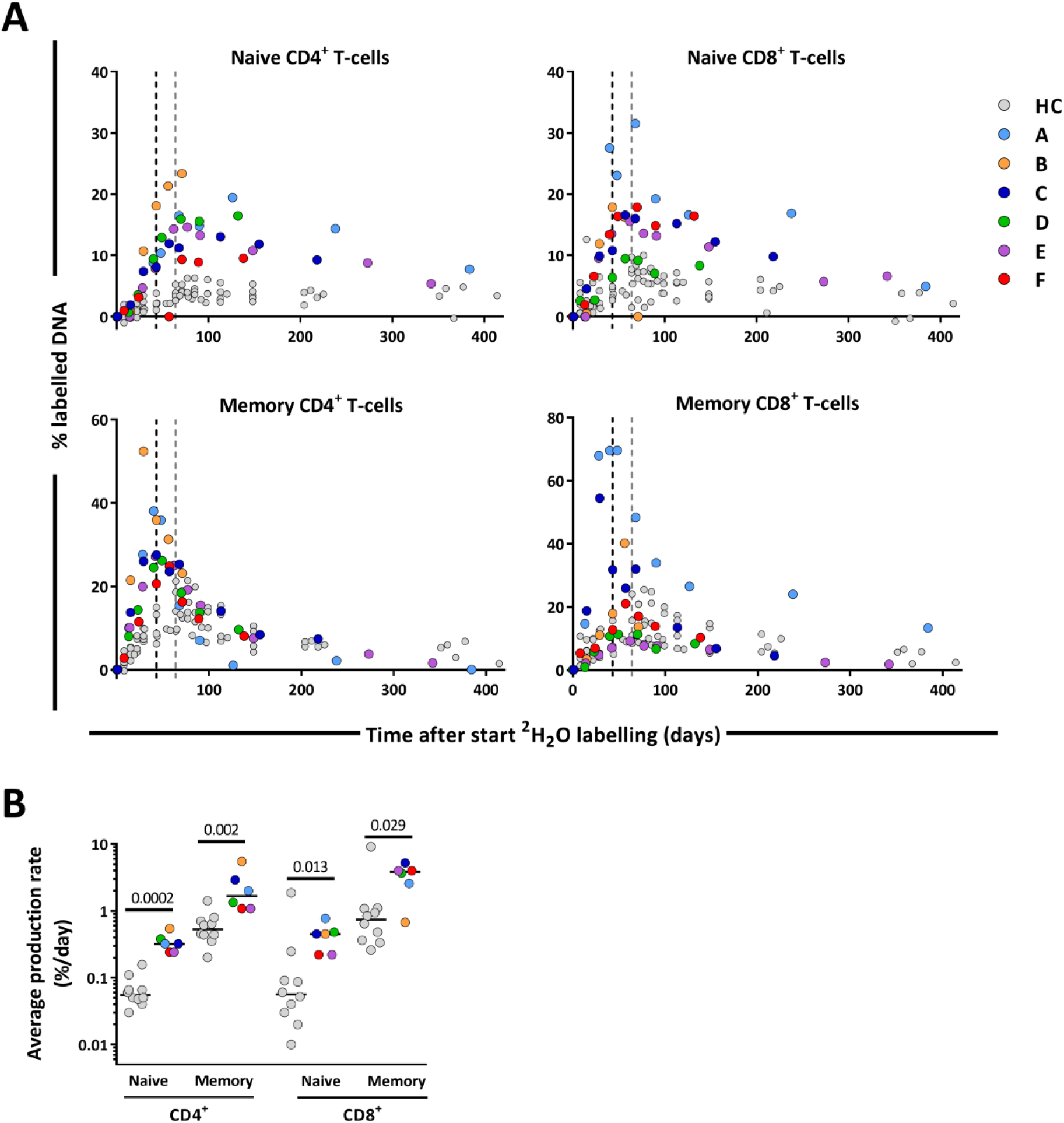
T-cell dynamics after autoHSCT. **(A)** Deuterium enrichment in the DNA of naive and memory CD4^+^ and CD8^+^ T-cells in autoHSCT patients (A to F, color symbols), and healthy controls (HCs, grey symbols) ^16^. Dotted lines correspond to the end of the labelling period (black for autoHSCT patients and grey for HCs). Label enrichment was scaled between 0 and 100% by normalizing for the maximum enrichment in granulocytes (Sup. Figure 7). For individual fits see Sup. Figure 8. **(B)** Estimates of the per cell production rate of naive and memory CD4^+^ and CD8^+^ T-cells in autoHSCT patients and HCs ^16^. Different symbols indicate different individuals, autoHSCT patients (A to F) in color and HCs in grey. Horizontal lines represent median values. P-values of significant differences between groups are shown.

Because it is generally assumed that during lymphopenia the availability of growth and survival factors increases, which has in particular been shown for IL-7 plasma levels ^18–22^, we also determined plasma levels of IL-7 and IL-15 between 12 and 24 months post-autoHSCT. Despite the CD4^+^ T-cell lymphopenia observed in these patients, their plasma concentrations of IL-7 and IL-15 and several other cytokines were in the range of those of healthy controls (Sup. Figure 2).

### Increased CD4^+^ and CD8^+^ T-cell production rates post-autoHSCT

To investigate whether low CD4^+^ T-cell numbers were associated with increased T-cell production rates, we compared the level of deuterium enrichment in the DNA of the different T-cell subsets between patients and controls. Deuterium enrichment analysis showed a relatively high level of label incorporation in patients, despite the fact that the labelling period was 3 weeks shorter for patients than for controls (Figure 3A). Using mathematical modelling we estimated the production rates of the different T-cell subsets (i.e. the number of new cells produced per day, coming from a source or peripheral cell division, divided by the number of resident cells in the population). We found that the production rates of naive and memory CD4^+^ T-cells were, respectively, 6-times and 3-times higher in patients than in controls. For naive and memory CD8^+^ T-cells, the estimated production rates were approximately 8- and 4-times higher in patients compared to controls (Figure 3B, Sup. Table 1), despite the fact that absolute CD8^+^ T-cell numbers had already recovered to healthy levels 12 months post-transplantation.

### Increased proliferation of naive but not memory CD4^+^ and CD8^+^ T cells

T-cell production rates as measured by deuterium labelling may reflect cell division of the subset of interest or an influx of cells from a source (e.g. by thymic output) or from another subset (e.g. through lymphocyte differentiation). To distinguish between these options, we first measured Ki-67 expression, a marker for peripheral cell division, of all lymphocyte subsets. The fraction of Ki-67^+^ cells within the naive CD4^+^ and CD8^+^ T-cell pools was significantly higher in patients compared to controls (Figure 4A). For the memory T-cell subsets, in contrast, the fraction of Ki-67^+^ cells of patients did not differ significantly from those of controls (Figure 4A). This suggests that the increased production rates of memory CD4^+^ and CD8^+^ T-cells may occur due to an increased influx from naive T-cells into the memory compartment, rather than increased T-cell division within the memory T-cell pools.

**Figure 4.**
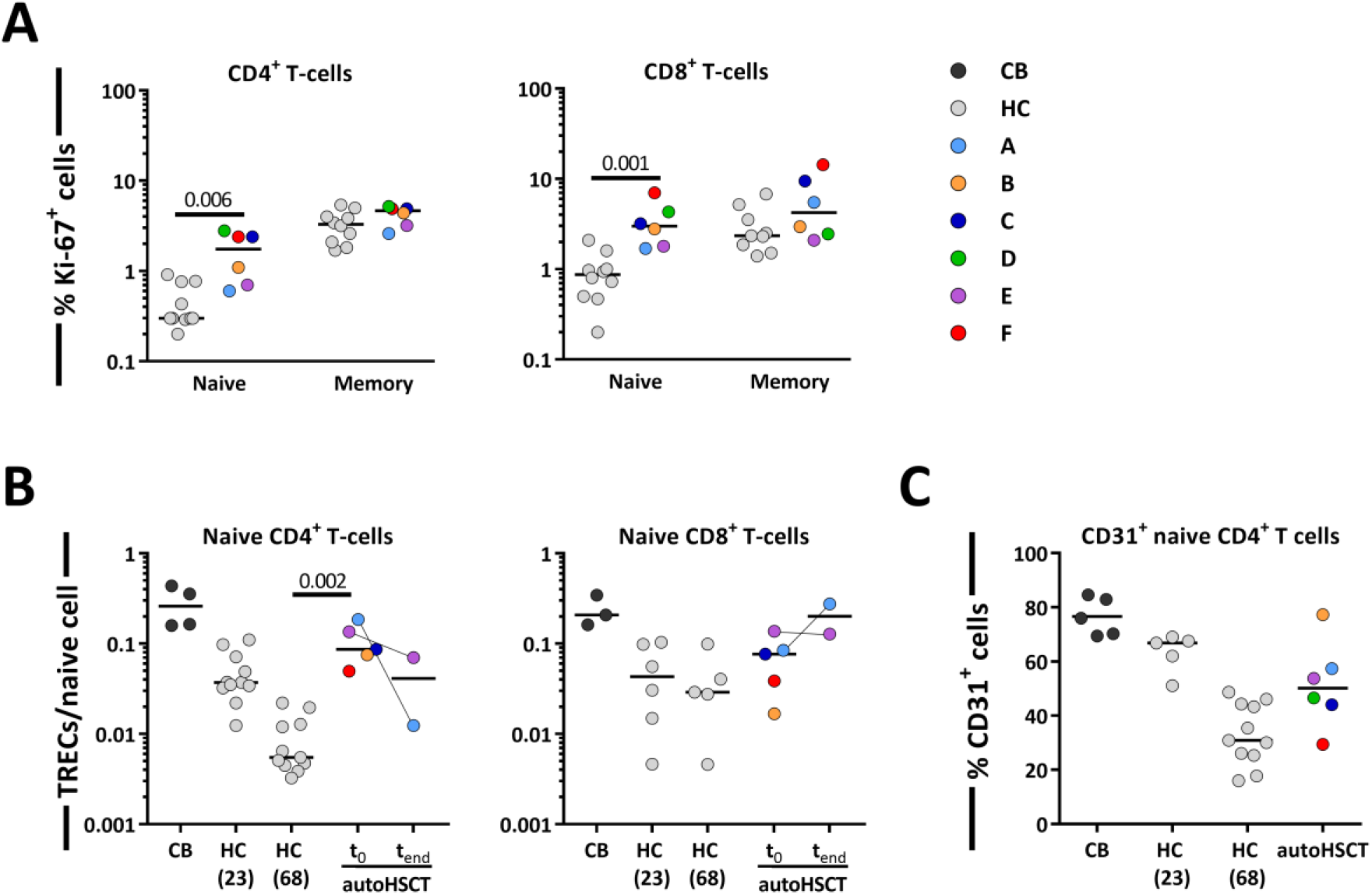
Contribution of peripheral proliferation and thymic output to T-cell production after autoHSCT. **(A)** Ki-67 expression was measured within naive and memory CD4^+^ (left panel) and CD8^+^ (right panel) T-cell in autoHSCT patients and HCs ^16^. **(B)** Average number of TRECs per naive CD4^+^ (left panel) and CD8^+^ (right panel) T-cell in autoHSCT patients, cord blood (CB) and healthy controls (HCs) ^16^. For *Patient A* and *Patient E*, TREC content was measured the first day of the study (t_0_) as well as the last study visit (t_end_). For *Patient D*, TREC content was not successfully measured due to limited material. **(C)** CD31 expression was measured within naive CD4^+^ T-cells in autoHSCT patients, cord blood and HCs ^16^. For changes in CD31 expression and absolute numbers of CD31^+^ cells over time, see Sup. Figure 4. Different symbols indicate different individuals, autoHSCT patients (A to F) in color, CB in dark grey, and young (median age of 23 years) and old (median age of 68 years) HCs in light grey. Horizontal lines represent median values. P-values of significant differences between groups are shown.

Besides increased cell division in the naive T-cell pool, increased naive T-cell production rates post-autoHSCT could in theory also be due to increased thymic output. T-cell receptor excision circles (TRECs) are commonly measured to estimate thymopoiesis. Because the average TREC content per T-cell declines with age ^23–25^, we measured TREC contents of naive T-cells from patients, cord blood, and young (on average 23 years of age) and aged (on average 68 years of age) healthy individuals ^16^. The average TREC content of naive CD4^+^ T-cells in patients was approximately 10-fold higher than in aged controls, and not significantly different from that of young individuals and cord blood (Figure 4B), even though all but one of the patients were more than 50 years of age. For naive CD8^+^ T-cells, the average TREC content in patients was in the range of young and aged controls (Figure 4B). We also measured CD31 expression on naive CD4^+^ T-cells, as CD31^+^CD4^+^ T cells are known to be enriched in recent thymic emigrants (RTEs) ^26,27^. The fraction of CD31^+^ cells within the naive CD4^+^ T-cell population was slightly higher in patients than in aged controls and slightly lower than in young controls and cord blood (Figure 4C, Sup. Figure 4). For naive T-cells, the combined Ki-67, TREC and CD31 data suggest that the increased T-cell production is due to increased T-cell division, and that the increased average TREC contents and percentages of CD31^+^ cells may be a direct consequence of normal thymic output entering a smaller T-cell pool ^28^.

### Heterogeneous B-cell reconstitution kinetics post-autoHSCT

Next, we studied the changes in B-cell dynamics following autoHSCT. Although total CD19^+^ B- cell numbers and naive (IgM^+^CD27^-^) B-cell numbers had already reached normal or even supra-normal levels by day 200 post-autoHSCT, Ig class-switched (IgM^-^CD27^+^) and IgM^+^ (IgM^+^CD27^+^) memory B-cell numbers in most patients were still below, or in the lower range of, those of healthy controls throughout the study period (Figure 5A, 5B and Sup. Figure 5).

**Figure 5.**
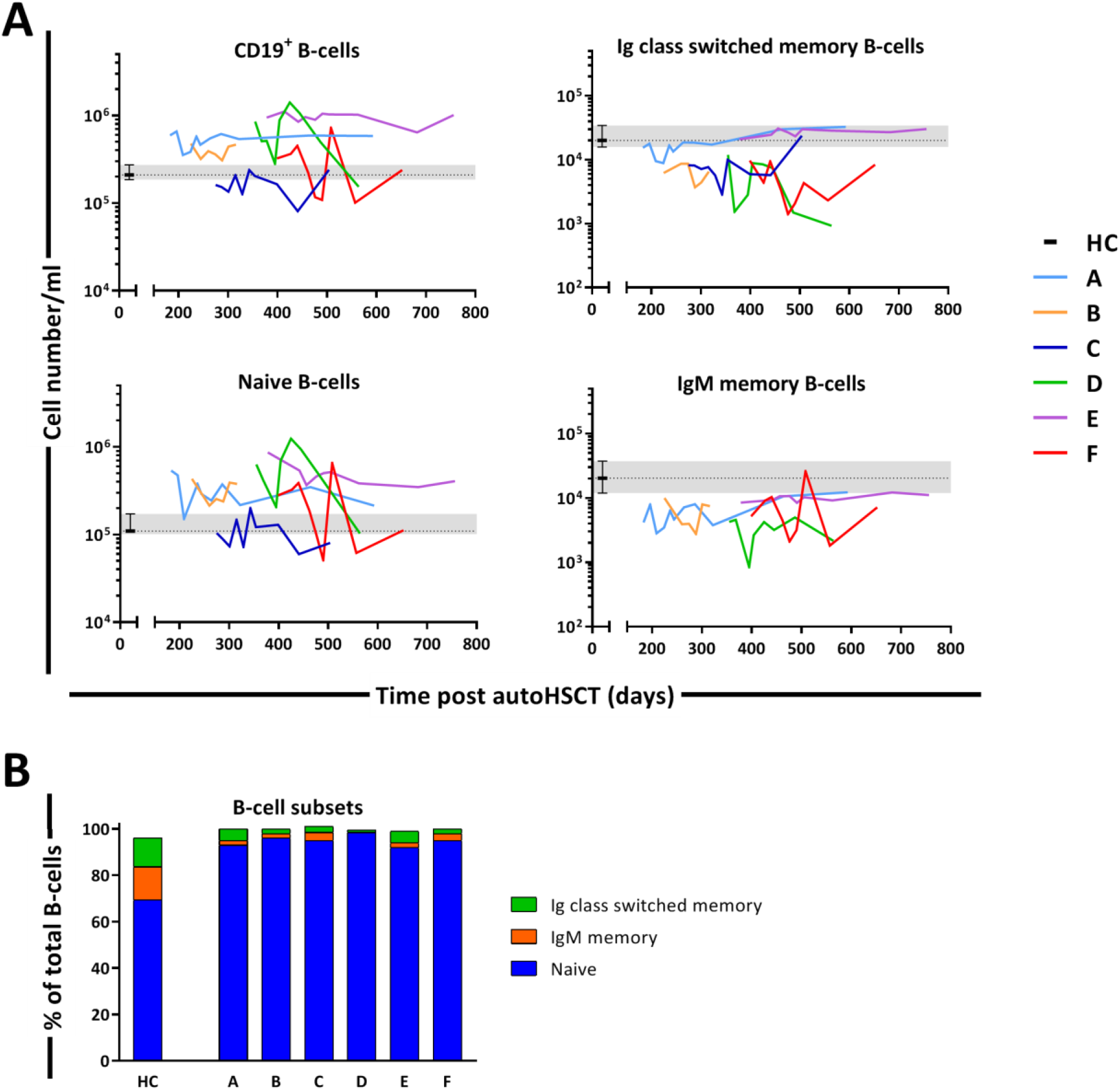
B-cell reconstitution following autoHSCT. **(A)** Absolute numbers (cells per milliliter) of total CD19^+^ B-cells, naive (CD19^+^IgM^+^CD27^-^), Ig class-switched memory (CD19^+^IgM^-^CD27^+^) and IgM^+^ memory (CD19^+^IgM^+^CD27^+^) B-cells in peripheral blood over time. Graphs show the absolute cell counts per milliliter in autoHSCT patients (patients A to F) over the duration of the study, and the median and the corresponding IR (interquartile rage) for HCs (healthy controls, grey area and whisker bars). **(B)** Bar graphs show the median percentage of naive, Ig class-switched memory and IgM^+^ memory B-cells within total CD19^+^ B-cells of autoHSCT patients (patients A to F) and HCs (naive in blue, natural effector in orange and memory in green). For the B-cell subset distribution per patient over time see Sup. Figure 5. Note the different y-axes in panel A.

### Increased production rates of B-cells post-autoHSCT

We analysed the deuterium enrichment of the different B-cell subsets to study whether B-cell production rates were increased for subsets which were still low in cell numbers (Figure 6A). The production rates of Ig switched-memory B-cells and IgM^+^ memory B-cells were 3.5-times and 5-times higher than in controls, respectively (Figure 6B, Sup. Table 1). Also the production rate of naive B-cells, a population that had already reconstituted to supra-normal levels, remained significantly higher than in healthy controls (Figure 6B, Sup. Table 1).

**Figure 6.**
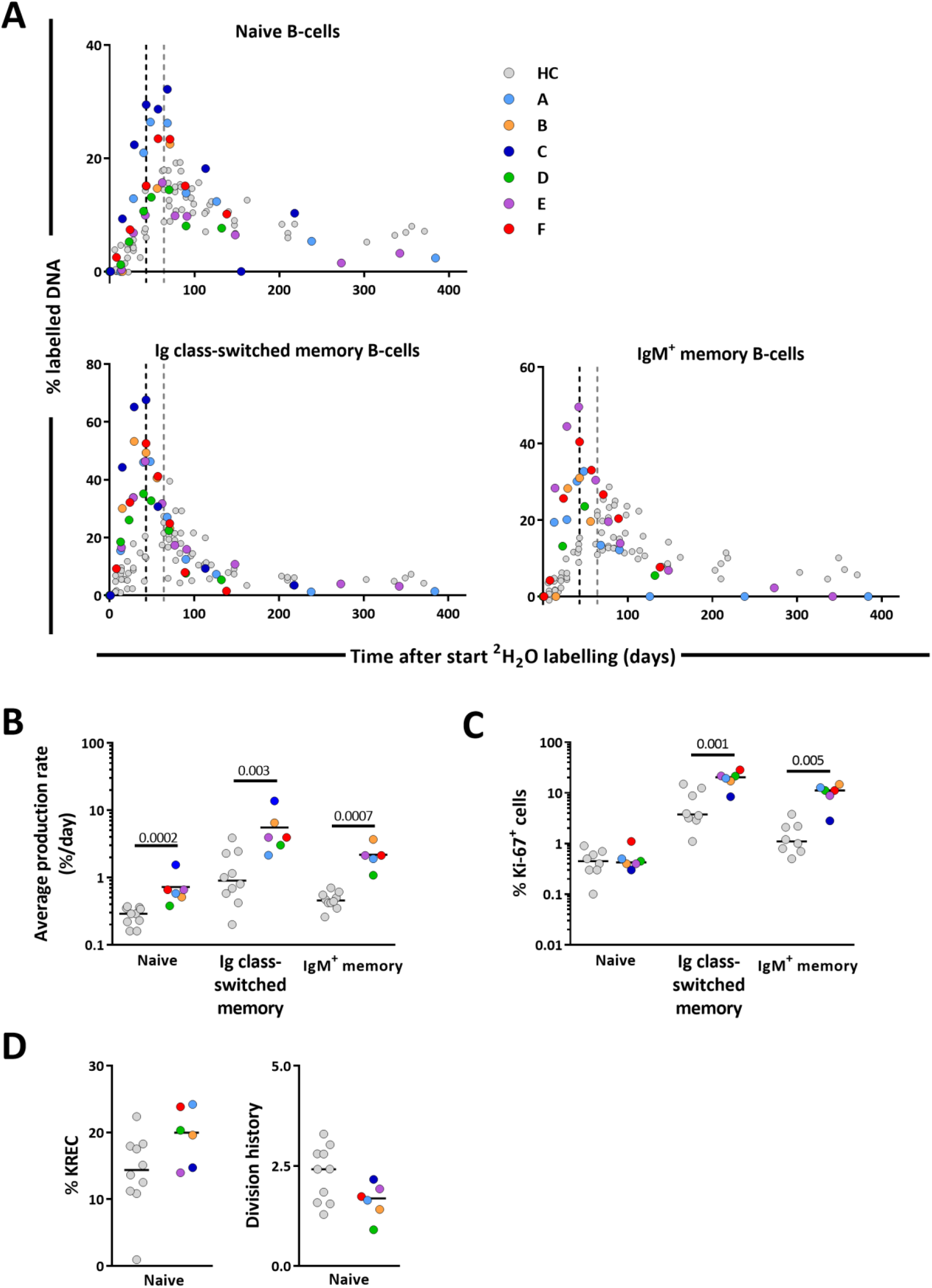
B-cell dynamics after autoHSCT. **(A)** Deuterium enrichment in the DNA of naive, Ig class-switched memory and IgM^+^ memory B-cells in autoHSCT patients (A to F, color symbols), and healthy controls (HCs, grey symbols) ^16^. Dotted lines correspond to the end of the labelling period (black for autoHSCT patients and grey for HCs). Label enrichment was scaled between 0 and 100% by normalizing for the maximum enrichment in granulocytes for each patient (Sup. Figure 8). For individual fits see Sup. Figure 9. **(B)** Estimates of the per cell production rates of naive, Ig class-switched memory and IgM^+^ memory B-cells in autoHSCT patients and HCs ^16^. **(C)** Ki-67 expression was measured within naive, Ig class-switched memory and IgM^+^ memory B-cells in autoHSCT patients and HCs ^16^. **(D)** Percentage of naive B-cells containing a KREC and naive B-cell replication history for autoHSCT patients and HCs ^16^. Different symbols indicate different individuals, autoHSCT patients (A to F) in color and HCs in grey. Horizontal lines represent median values. P-values of significant differences between groups are shown.

Because B-cell production may depend on peripheral B-cell division and on *de novo* bone marrow output, we measured Ki-67 expression and kappa-deleting recombination excision circles (KRECs), in an attempt to estimate bone marrow output. The percentages of dividing, i.e. Ki-67^+^, cells within IgM^+^ and Ig switched-memory B-cells were significantly higher in patients than in healthy individuals (Figure 6C). In contrast, the fraction of Ki-67^+^ cells within the naive B-cell subset was similar between patients and controls (Figure 6C). Although naive B-cell peripheral division rates were not increased post-autoHSCT, their production rates were 2-times higher than in controls. The replication history (measured as number of cell divisions) of the naive B-cell subsets in patients tended to be lower than in controls (although not significantly), suggesting that the output of naive B-cells from the bone marrow rather than their peripheral proliferation rate was increased after autoHSCT (Figure 6D).

### Increased lymphocyte production rates are counteracted by increased lymphocyte loss rates

The increased lymphocyte production rates that we observed in patients after autoHSCT may at first sight suggest that, also in humans, lymphocyte production is regulated in a density-dependent manner. The observation that lymphocyte production rates were also elevated for subsets for which cell numbers had already normalized, however, challenges this interpretation. Another observation challenging this interpretation is that for most subsets, lymphocyte numbers increased very little over time, despite the significant increase in lymphocyte production. This suggests that lymphocyte loss rates were also significantly increased after autoHSCT.

To estimate the average loss rates of all lymphocyte subsets (i.e. the number of cells lost per day, by cell death, migration or differentiation, divided by the number of resident cells in the population), we used the average lymphocyte production rates estimated from the deuterium labelling experiments and an exponential function to describe the changes in cell numbers of each lymphocyte subset over time (see Sup. material and methods). For most T-cell and B-cell subsets the average loss rate was approximately 3 to 5-times higher post-autoHSCT than in healthy individuals (Figure 7, Sup. Table 2). For naive CD8^+^ T-cells the average loss rate was even 9.5-times higher in patients than in healthy individuals (Figure 7, Sup. Table 2). Thus, despite the fact that production rates are clearly increased in patients post-autoHSCT, this increased production goes hand in hand with increased lymphocyte loss rates, thereby challenging the view that it reflects a homeostatic response to low lymphocyte numbers.

**Figure 7.**
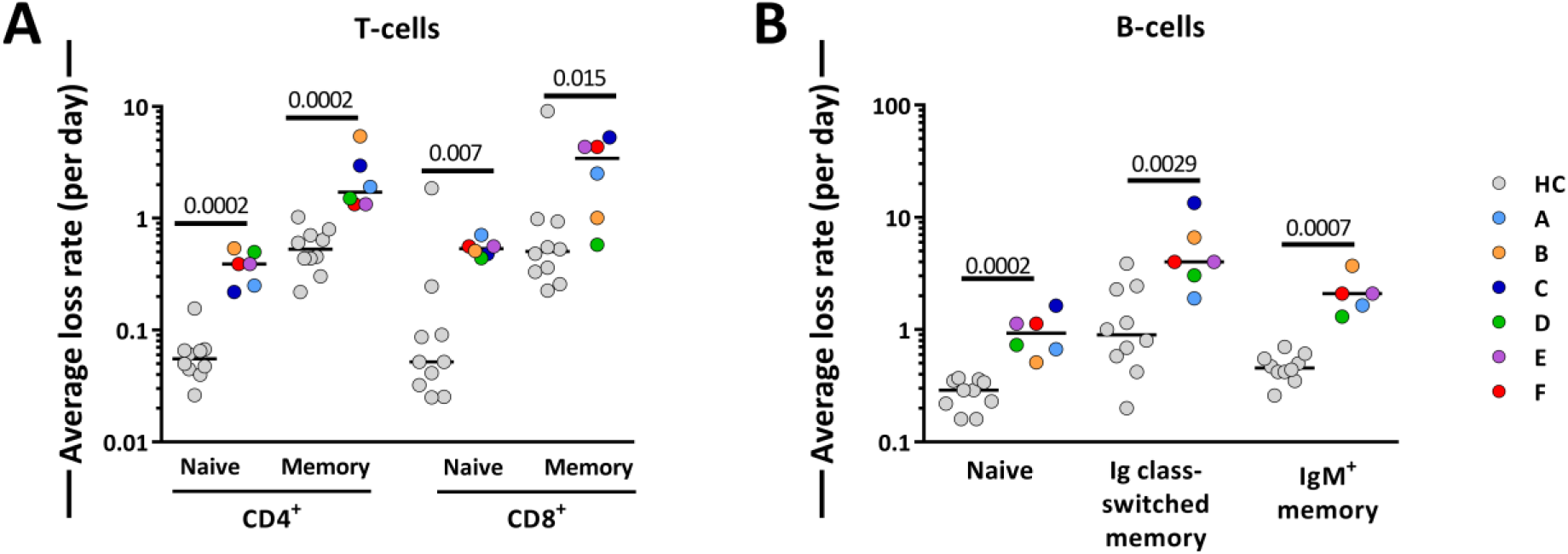
Average T-cell and B-cell loss rates following autoHSCT. **(A)** Estimates of the average loss rates of naive and memory CD4^+^ and CD8^+^ T-cells and of **(B)** naive, Ig class-switched memory and IgM^+^ memory B-cells in autoHSCT patients (A to F, color symbols), and healthy controls (HC, grey symbols) ^16^. Average loss rates were calculated using the estimated average turnover rates and the corrected cell numbers (Sup. Figure 10 and 11) as described in supplementary material and methods. Horizontal lines represent median values. P-values of significant differences between groups are shown.

## Discussion

From a homeostatic viewpoint, a response to low lymphocyte numbers could take the form of increased lymphocyte production or decreased lymphocyte loss. Based on the observation that severe lymphopenia in mice is associated with increased peripheral proliferation ^3–5,20^, it is widely believed that lymphocyte production rates are increased when cell numbers are low. We have previously shown that naive T-cell turnover rates do not increase to compensate for the at least 10-fold decline in thymic output in elderly individuals ^16^. This could be due to the relatively small degree of naive T-cell loss observed during healthy ageing. Under more severe conditions of lymphopenia in humans, high frequencies of proliferating lymphocytes have been observed, but these have been linked to immune activation and clinical events, e.g. GVHD and opportunistic infections ^14^. Thus, there is no convincing evidence that homeostatic regulation of lymphocyte production and loss occurs in humans.

Our deuterium labelling study shows that in patients receiving an autoHSCT, in the absence of GVHD, clinically manifested infections and transplantation-related complications, the production rates of most T- and B-cell subsets were significantly increased 12 months after transplantation. Increased lymphocyte production following lymphodepletion has generally been interpreted as evidence for a density-dependent response to low lymphocyte numbers ^12,13,15,29,30^. Our study challenges this view for two reasons: i) T- and B-cell production rates did not normalize when cell numbers did, and ii) lymphocyte loss rates were also increased post-autoHSCT. This suggests that the increase in lymphocyte production rates in autoHSCT patients was not simply a homeostatic response to low cell- numbers. Other determinants, such as repertoire diversity or the clonality of the T-cell pool might also play a role. In addition, despite the fact that the patients in our study were included up to 12 months post-transplantation and were selected on the basis of being in good health, we cannot exclude that the increased rates of lymphocyte production and loss in these patients reflect HSCT-related complications, such as the impact of initial chemo-therapy and conditioning-therapy or sub-clinical infections and inflammation, which may have gone unnoticed.

The observation that lymphocyte loss rates were increased post-autoHSCT is remarkable in the light of the widely held view that homeostatic mechanisms could take the form of increased lymphocyte survival. This concept is supported by the observation that the availability of pro-survival and anti-apoptotic factors, such as IL-7 ^31^, increases during lymphopenia. We found that lymphocyte loss rates were up to 10-fold increased after autoHSCT. This is in line with previous human studies on T-cell survival after allogenic HSCT, which consistently reported that the fraction of pro-apoptotic cells increases following transplantation ^15,32–34^. Although this suggests that intervention with lymphocyte survival after HSCT may aid lymphocyte reconstitution, in our study, different factors may have contributed to the loss of cells from the peripheral blood. Cells may have been lost from the circulation by cell death, but also by lymphocyte differentiation and/or migration to the tissues. Further studies need to clarify whether lymphocyte reconstitution occurs at similar rates in blood and tissues, or whether lymphocyte recruitment to the tissues may be a key factor influencing the loss of lymphocytes from the blood following autoHSCT.

Consistent with previous reports ^10,15,35^, we found that 12 months post-autoHSCT, CD4^+^ T-cell numbers were below the normal range while CD8^+^ T-cells recovered more rapidly. Deuterium labelling in patients revealed that the average production rates of most T-cell subsets were significantly increased following autoHSCT. This increase was especially evident for naive T-cells. The high percentage of Ki-67^+^ naive T-cells post-autoHSCT suggests that increased naive T-cell production is to a large extent explained by increased peripheral T-cell proliferation. Memory T-cell production rates (based on deuterium enrichment) were also higher in patients compared to controls, while Ki-67 expression suggested that memory CD4^+^ and CD8^+^ T-cell proliferation rates were not increased after autoHSCT. This seeming contradiction may be explained by the fact that Ki-67, a snapshot marker, may be less sensitive to detect differences in T-cell proliferation than long-term *in vivo* deuterium labelling. Alternatively, the increased production rate of memory T cells post-autoHSCT may be due to increased transition of naive T cells into the memory T-cell population. In mice it has been demonstrated that naive T-cells adoptively transferred into immunodeficient animals can undergo division without cognate antigen stimulation, thereby acquiring a memory phenotype ^36,37^.

If a significant part of cell production in a certain lymphocyte subset (e.g. the memory subset) is indeed due to an influx from another lymphocyte subset (e.g. the naive subset), the increased production rates that we observed may either reflect a true increase in cell production, or a normal influx of cells entering a smaller lymphocyte population. To distinguish between these options, for each lymphocyte subset and each individual, we also calculated the total number of cells produced per day (i.e. coming from a source and/or from peripheral cell division), by multiplying the average production rate of each lymphocyte subset with the median cell number of that subset, and compared these values to those in healthy controls (data not shown). We found that total daily lymphocyte production was as high as in healthy controls for naive CD4^+^ T cells and higher than in healthy controls for all other lymphocyte subsets, suggesting that the increased lymphocyte production rates post-autoHSCT truly reflected increased T-cell proliferation and/or an increased influx from another lymphocyte compartment.

Measuring thymopoiesis and the contribution of recent thymic emigrants to the naive T-cell pool after HSCT is not straightforward. Although increased TREC contents at first sight seem suggestive for increased thymic output, T-cells bearing TRECs may in fact be overrepresented in the peripheral T-cell pool post-transplantation when cell numbers are low ^28^. Hence, for naive CD4^+^ T cells, whose numbers had not yet normalized, increased average TREC contents may incorrectly be interpreted as evidence for increased thymic output. The finding that the average TREC content of naive CD4^+^ T-cells following autoHSCT was higher than in age-matched controls provides no evidence that thymic output following transplantation was higher than in healthy controls, but does imply that the thymus had become functional again within 12 months after intense conditioning for autoHSCT. The fact that the average TREC content of naive CD4^+^ T -cells, but not that of naive CD8^+^ T-cells, was higher in patients than in healthy individuals may reflect differences in the degree of depletion of naive CD4^+^ and CD8^+^ T -cells. Alternatively, it might reflect differences in the way CD4^+^ and CD8^+^ T-cells are generated. In support of the latter explanation, repertoire analyses in patients receiving an autoHSCT for the treatment of autoimmune diseases have suggested that CD4^+^ T-cells largely arise *de novo*, since most CD4^+^ T-cell clones post-autoHSCT were not present at baseline, while CD8^+^ T-cells mainly expand from cells that were already circulating pre-transplantation ^38–40^.

To study in a population other than T-cells whether lymphocyte production and loss rates in humans are regulated in a density-dependent manner, we quantified the production and loss rates of different B-cell subsets. In line with previous reports ^41–43^, we found that 12 months after transplantation, naive B-cell numbers had reconstituted to healthy (or even higher than healthy) control values, while Ig class-switched and IgM^+^ memory B-cells had not yet fully recovered. The delayed reconstitution of Ig class-switched and IgM^+^ memory B-cells has typically been attributed to treatment-related damage to secondary lymphoid organs, which may hamper the formation of germinal centers essential for somatic hypermutation and isotype switching ^42^. Also for naive, Ig class-switched, and IgM^+^ memory B-cells, we found that not only production rates but also cell loss rates were increased 12 months post-autoHSCT, further supporting our conclusion that increased lymphocyte production rates do not simply reflect a homeostatic response to low lymphocyte numbers.

In brief, our findings show that despite the slow reconstitution of lymphocytes in autoHSCT patients, lymphocyte production rates are increased is. Since this increased production goes hand in hand with increased cell loss, and does not normalize when cell numbers do, it is not simply due to homeostatic mechanisms. Future studies should address whether the dynamics of lymphocytes after autoHSCT normalize in the long run, what drives the increase in lymphocyte production and loss rates during immune reconstitution, and to what extent immune reconstitution in the tissues occurs.

## Data Availability

Data will be available upon request to the corresponding author.

## Acknowledgments

We thank the patients for their participation in this study, Jeroen F. van Velzen, Pien A.J. van der Burght and Gerrit Spierenburg for assistance with cell sorting, Laura Ackermans for theoretical input, Lyanne Derksen for critically reading the manuscript, Mr Benjamin Bartol and Ms Pei Mun Aui for technical support, and the nurses from the Julius Center Trial Unit for taking care of study participants.

## Authorship

Contribution: MB-P, VvH, JD, JB and KT designed the experiments; MB-P and VvH performed the experiments; LvdW and AJ selected and included the patients; MB-P, VH, JD, RdB, JB and KT analysed and interpreted data; JD, RdB, and JB performed mathematical modelling; and all authors wrote and approved the manuscript.

